# Doctors’ experiences of interruptions in UK secondary care outpatient clinics: a cross-sectional survey of prevalence, consequences and management strategies

**DOI:** 10.64898/2026.10.07.26364858

**Authors:** Shaun Boustani, Chase Staras, Antonia Rich, Ann Griffin

**Author notes:** **Corresponding author:** Dr Shaun Boustani.

## Abstract

**Objectives:** To examine the prevalence of interruptions during UK secondary care outpatient consultations, their association with clinician mental workload, and the strategies clinicians use to manage them.

**Design:** Nationwide cross-sectional online survey.

**Setting:** Secondary care outpatient clinics, United Kingdom.

**Participants:** A sample of 465 UK secondary care doctors across a range of grades and specialties, recruited through a medical defence organisation’s network.

**Primary and secondary outcome measures:** Mental workload (NASA Task Load Index, Raw-TLX); interruption frequency; perceived cognitive impact of interruptions; task resumption difficulty; perceived social effects on colleague interactions and team cohesion; and interruption-management strategy use.

**Results:** 61.1% of clinicians reported being interrupted often or very often, and mean overall workload was high (Raw-TLX 66.2, SD 11.5). Interruption frequency, patient complexity and perceived cognitive impact were each associated with higher workload across multiple NASA-TLX subscales. In fully adjusted models, interruption frequency retained independent associations with overall workload (β = 1.79, p = 0.004) and temporal demand (β = 0.94, p < 0.001). Task resumption difficulty was associated with worse perceived performance (β = −0.59, p = 0.014), while perceiving interruptions as socially positive was associated with better performance (β = 0.75, p = 0.003) and lower frustration (β = −1.16, p < 0.001). Years of experience showed only small effects, and self-reported strategy use (38.7%) was not consistently associated with lower workload. Exploratory natural language processing of open-text descriptions identified three strategy clusters; cognitive offloading, anticipating and preparing, and blunt responses, with distinct emotional profiles.

**Conclusions:** Interruptions were commonly reported during UK secondary care outpatient consultations and were associated with measurable cognitive and temporal burden. The perceived social framing of interruptions was independently associated with frustration and self-rated performance. Binary strategy use showed no consistent workload association, indicating a need for system-level approaches addressing interruption frequency, task resumption support and the social context of interruptions rather than reliance on individual coping.

**Strengths and limitations of this study:**

- To our knowledge, this is the first national survey of interruptions in UK secondary care outpatient consultations, spanning a wide range of specialties, grades and career stages (0–52 years post-qualification).
- Mental workload was measured using the NASA Task Load Index, a validated multidimensional instrument, allowing workload to be decomposed into cognitive, temporal, physical and affective components.
- Open-text descriptions of interruption-management strategies were analysed using natural language processing, revealing heterogeneity in strategy type that binary coding alone would have obscured.
- The cross-sectional design precludes causal inference, and the direction of the association between perceived interruption frequency and workload cannot be established.
- Recruitment through a single medical defence organisation’s network, together with an indeterminate eligible denominator, means this was a convenience sample in which non-response bias cannot be excluded.

## Introduction

Work interruptions are defined as “an unexpected suspension of the behavioral performance of, and/or attentional focus from, an ongoing work task” [1]. In healthcare, where complex clinical decisions must be made efficiently, accurately, and under time pressure, the consequences of interruptions extend beyond inconvenience. Research in cognitive psychology has established that interruptions increase the burden on memory and attentional resources, interfere with goal attainment, and elevate work-related stress [2]. However, research on the impact of interruptions during clinical consultations is limited. Clinical consultations are the core interaction by which secondary care clinicians diagnose and manage patients, and safeguarding their quality is central to patient safety and healthcare effectiveness [3]. The scale of outpatient activity amplifies this concern. In England alone, there were 104.6 million outpatient appointments in 2023–24, making the secondary care outpatient clinic one of the most consequential clinical settings in the health system [4]. This study aims to fill this gap by investigating clinicians’ perceptions of interruption frequency, the perceived cognitive and operational impacts of interruptions, and the contextual and social factors that shape how interruptions are experienced and managed during secondary care consultations.

### Cognitive load and the impact of interruptions

The impact of interruptions on clinical performance can be understood through Cognitive Load Theory (CLT), which holds that working memory has a finite capacity, and that additional demands imposed by interruptions (which can be considered an extraneous load) disrupt task continuity and reduce the efficiency of cognitive processing [5]. When an interruption occurs, clinicians must shift attentional focus, reallocate cognitive resources, and subsequently re-engage with the interrupted task, each of which imposes additional cognitive costs. Repeated interruptions throughout a clinic session can compound these costs, culminating in cognitive fatigue [6,7].

However, the degree to which an interruption is disruptive is not uniform. CLT identifies task complexity as a key moderating variable. Tasks characterised by a higher number of interrelated elements, greater uncertainty, and more demanding decision-making have greater intrinsic cognitive load, and are therefore more susceptible to the disruptive effects of an interruption [8,6]. In the context of outpatient secondary care, patient complexity serves as a meaningful index of this intrinsic task demand. Patient complexity reflects the interplay of medical and non-medical factors, including comorbidities, polypharmacy, mental health conditions, and socioeconomic challenges, that challenge routine clinical management [9]. As patient complexity increases, clinicians must integrate more information, manage greater uncertainty, and make decisions under time pressure, thereby elevating the effective cognitive load of the clinical encounter [10].

Conversely, CLT also proposes that clinician experience provides a partial buffer against this burden. Experienced practitioners will have developed automated routines and well-organised mental frameworks that allow them to process familiar clinical information more efficiently, a phenomenon described by the expertise reversal effect [11]. Novices, lacking these established schemas, are likely to experience comparatively greater cognitive strain, particularly when confronted with complex patients or frequent interruptions. Despite this, little research has consistently demonstrated that experience confers meaningful protection against interruption-related workload, and it remains unclear whether this holds in secondary care outpatient contexts.

### Interruptions in healthcare settings

In a systematic review, Rivera-Rodriguez and Karsh [12] identified that interruptions in healthcare settings are common, particularly in hospital wards. Although evidence from outpatient settings is more limited, available studies suggest they are similarly prevalent. For example, in one Israeli general practice study, 94% of consultations were interrupted [13], and qualitative data from general practice indicate that interruptions are perceived negatively by clinicians and can undermine both consultation quality and the doctor–patient relationship [14,15]. Interruptions at critical junctures, such as during medication prescribing or procedural steps, carry implications for patient safety [16,3] and have been shown to unnecessarily extend consultation length, exacerbating existing pressures on waiting lists [17]. Despite the scale of outpatient activity and the established cognitive costs of interruptions, relatively little is known about how frequently secondary care clinicians experience interruptions, the magnitude of their effect on perceived workload, and whether clinicians employ strategies to manage them.

### Social dimensions of interruptions

Interruptions are not solely cognitive events; they are inherently interpersonal. An interruption is an interaction between an interrupter and an interrupted individual, and as such is embedded within a team’s dynamics and shaped by interprofessional communication norms [18]. Research from organisational psychology has demonstrated that the social consequences of interruptions are dual-natured. While interruptions can derail collaboration, trigger negative commentary, and erode team cohesion, they can also catalyse constructive exchanges, including humour, encouragement, and supportive communication, that reaffirm team bonds [18,19]. Puranik et al. [20] found that, while the task-switching demands imposed by interruptions deplete self-regulatory resources and undermine well-being, the interpersonal interactions inherent to them can simultaneously fulfil fundamental belongingness needs, partially offsetting the negative effects. Brief, courteous exchanges during an interruption, such as an apology or an expression of acknowledgement, may help mitigate cognitive costs by sustaining positive affect and social connection.

This social dimension is particularly salient in outpatient secondary care, where clinicians may be interrupted by a range of individuals, including colleagues, trainees, administrative staff, and other patients. Whether an interruption is managed constructively or abruptly has implications not only for the clinician’s immediate cognitive state but also for the interprofessional relationships and team cohesion that underpin safe and effective care. Research on structured interprofessional communication approaches, such as the Identification–Situation–Background–Assessment–Recommendation (ISBAR) handover framework [21], suggests that systematic protocols can improve the quality of task handovers and reduce cognitive disruption. However, whether analogous strategies benefit clinicians managing consultation interruptions remains underexplored.

### Management of interruptions

Although clinical interruptions are widely acknowledged as problematic, evidence on effective management strategies remains limited. Attempts to reduce interruption frequency through intervention have yielded mixed results [12,22], and complete elimination is neither realistic nor always desirable, since some interruptions convey time-critical or clinically essential information. What remains largely unknown is whether and how clinicians actively manage interruptions in practice, what strategies they employ, and whether such strategies are associated with meaningfully lower cognitive workload. A binary view of “strategy use” is unlikely to be informative; the type, fidelity, and contextual fit of a strategy may matter more than its mere presence [20].

### The present study

The present study addresses these gaps using a cross-sectional survey of secondary care clinicians in the United Kingdom (UK). We use the NASA Task Load Index (Raw-TLX) [23], a validated multidimensional instrument adapted for healthcare contexts [24,25,26], to examine levels of clinician mental workload and explore their associations with interruption frequency, patient complexity, experience, and the perceived cognitive and social consequences of interruptions. We also examined the interruption-management strategies clinicians report using, characterising them using natural language processing (NLP) methods to move beyond binary reporting and identify the types, emotional profiles, and structural patterns of strategies in use.

Grounded in Cognitive Load Theory and perspectives on social dynamics, we anticipated that interruption frequency and patient complexity would independently predict higher workload, and that the perceived cognitive impact of interruptions would account for a substantial share of the association between frequency and workload. We further expected that perceived social effects of interruptions, both the immediate momentary costs and those accruing across clinical sessions, would account for meaningful variance in frustration and perceived performance. We also explored whether years of clinical experience influenced these associations, and whether strategy use was associated with lower workload, while remaining open to the possibility that these relationships are more complex than a simple buffering effect.

## Methods

### Design

This study employed a cross-sectional online survey administered using REDCap (Research Electronic Data Capture) [27]. This enabled a large-scale, systematic assessment of interruption frequency, perceived workload, and interruption-management strategies across a diverse sample of secondary care clinicians. Reporting follows the STROBE statement for cross-sectional studies and the Checklist for Reporting Results of Internet E-Surveys (CHERRIES).

### Sampling and recruitment

Recruitment was conducted in partnership with the Medical Protection Society (MPS) Foundation, the charitable research arm of a UK medical defence organisation. An email invitation describing the study was distributed on 14 July 2025 to the wider MPS network of approximately 73,787 UK doctors, with two follow-up reminders at two-week intervals (28 July and 11 August 2025). The survey closed on 18 August 2025. The invitation email was opened by 1,091 doctors, giving an indicative open rate of 1.5%.

This list is not a sampling frame of eligible clinicians: it included doctors not currently practising, those working exclusively in primary care, and those whose email engagement with the organisation was unknown. The true number of eligible doctors (currently working in UK secondary care outpatient clinics and who opened the recruitment email) cannot be reliably determined. A total of 465 eligible clinicians began the survey, yielding a crude participation rate of 42.6% among those who opened the invitation. The resulting sample was heterogeneous across specialty, grade, and career stage (0–52 years post-qualification), supporting broad relevance even if formal representativeness against the UK consultant workforce cannot be established. Non-response bias cannot be ruled out, and we discuss its likely direction in the Limitations.

No formal sample size calculation was performed; this was a convenience sample and all eligible respondents were included.

Because required-field validation was enabled, missing data reflect progressive drop-out across the instrument rather than item-level non-response. Of the 465 clinicians who began the survey, 430 (92.5%) reported a professional role, 429 (92.3%) reported a specialty, 415 (89.2%) completed the patient complexity item, 400 (86.0%) completed the NASA-TLX workload inventory, 391 (84.1%) completed the interruptions section, and 378 (81.3%) completed the teamwork items. Denominators are reported alongside each result throughout.

### Ethics and data governance

Ethical approval was obtained from the UCL Research Ethics Committee prior to data collection (ID: 1132; approved 20 May 2025). All participants completed an electronic consent form before accessing the survey. Consent records, which included the participant’s name and signature and optional contact details for future studies, were held separately from survey responses; analyses were conducted on a de-identified dataset in which responses were linked only to an automatically generated record identifier. Data are held within the UCL Data Safe Haven, accessible only to the named research team, in accordance with the terms of the ethical approval, and will be retained for no more than 10 years.

### Questionnaire development and pre-testing

The instrument comprised 34 fields presented across seven titled sections: 13 consent and administrative fields, 18 substantive research items, and 3 fields used to recruit to a planned focus-group study. The 18 analysed items are reproduced in full, with their response options, in the Supplementary Materials. It was reviewed by clinicians, educationalists and researchers at a departmental workshop and locally piloted prior to distribution, to sense-check item wording and response options and to test the usability and technical functionality of the electronic questionnaire. Items were presented in a fixed (non-randomised) order, and participants could review and change their responses using a back button. Two items were displayed conditionally: the open-text description of interruption-management strategies, shown only to participants reporting strategy use, and an email field used to recruit to a planned focus-group study. Required-field validation was enabled, so displayed items had to be answered before a participant could advance. No incentive was offered for participation.

### Measures

A detailed description of the measures used in this study can be found in the Supplementary Materials. In brief, patient complexity was assessed with a single item on a five-point scale, used as a proxy for intrinsic task demand consistent with cognitive load frameworks [8,9]. Mental workload was assessed using the NASA Task Load Index (NASA-TLX) [23], adapted for healthcare settings, yielding a Raw-TLX composite score (0–100) and six subscales: Mental Demand, Physical Demand, Temporal Demand, Performance, Effort, and Frustration. Interruption frequency, perceived cognitive impact of interruptions, and task resumption difficulty were each assessed with single Likert items. For the social effects of interruptions, no validated measure exists for clinical settings; items were therefore developed for this study, capturing two theoretically distinct dimensions, immediate interpersonal impact and broader team cohesion, drawing on Puranik et al. [1,20] and Toebben et al. [19]. Interruption-management strategy use was assessed as a binary variable (yes/no), with an open-ended field for respondents to describe the strategy used.

### Data analysis

#### Quantitative analyses

Data were analysed in R (version 4.5.0). Descriptive statistics (means, standard deviations, medians, frequencies, and proportions) were computed for all key variables. Bivariate associations between NASA-TLX outcomes and predictors (interruption frequency, patient complexity, perceived cognitive impact, task resumption difficulty, perceived social effects, strategy use, and years of experience) were examined using Pearson correlation coefficients (*r*), computed pairwise using all available data. Two-tailed *p*-values are reported, with an alpha level of .05. Effect sizes were interpreted using conventional benchmarks (small = .10, medium = .30, large = .50) [28].

For the primary analyses we required complete NASA-TLX data for a participant (all six subscales non-missing) before computing an overall Raw-TLX score; participants with any missing subscale were excluded from the overall score but were retained in item-level summaries to preserve power. Item-level summaries used all available data for each subscale.

To examine the independent contribution of interruption frequency to workload outcomes, sequential ordinary least-squares (OLS) regression analyses were conducted for each NASA-TLX outcome (overall Raw-TLX and each of the six subscales). For each outcome, five nested models were estimated on the same complete-case sample: Model 1 included interruption frequency only; Model 2 added patient complexity and years of experience; Model 3 added perceived cognitive impact and task resumption difficulty; Model 4 added perceived social effects of interruptions; and Model 5 added strategy use (binary). Unstandardised regression coefficients (*β*), standard errors (SE), and model *R*² are reported at each step.

#### Natural language processing of strategy descriptions

Open-text strategy descriptions were analysed using natural language processing (NLP) methods in R, using the tidytext and stringr packages. In brief, we used computational text-analysis methods to count recurring words and phrases, classify the emotional tone of each response, and group responses with similar wording into clusters. Text was pre-processed through standardisation, tokenisation into words and *n*-grams, and removal of common stop words. Word frequency analysis quantified the occurrence of individual words and higher-order *n*-grams (bigrams, trigrams, and six-grams) to identify recurring terms and phrases. Sentiment analysis was performed using the Bing Lexicon [29], with simple negations accounted for, and the NRC Emotion Lexicon [30] was applied to classify responses according to eight basic emotions (anger, anticipation, disgust, fear, joy, sadness, surprise, and trust) [31]. Semantic clustering used TF-IDF weighting and GloVe word embeddings to generate vector representations, followed by *k*-means clustering, with the final three-cluster solution selected on the basis of theoretical coherence and manual review. The emotional profile of each cluster was compared using chi-squared tests. Given the relatively small corpus of text (n = 145), the NLP and *k*-means clustering results are intended as an exploratory, proof-of-concept classification of strategic orientations. These computational clusters were manually reviewed to ensure thematic coherence and grounding in the raw data.

### Patient and public involvement

No patients or members of the public were involved in the design, conduct, reporting or dissemination plans of this research.

## Results

### Descriptives

A total of 465 clinicians began the survey; completion of individual components is reported alongside each result below. The mean time since qualification was 18.9 years (median = 19.0, range 0–52 years), indicating a predominance of mid- to late-career clinicians. Respondents came from a variety of professional roles (Table 1) and specialties (Table 2; full sub-specialty breakdown in Table S1).

**Table 1.** Professional roles of survey respondents (n = 430; 35 respondents did not report a role).

| Professional role | n | % |
| --- | --- | --- |
| Consultant | 326 | 75.8 |
| Specialty Registrar (SpR) | 27 | 6.3 |
| Specialty Trainee (ST) | 23 | 5.3 |
| Specialty Doctor (SAS) | 21 | 4.9 |
| Associate Specialist (SAS) | 9 | 2.1 |
| Senior House Officer (SHO) | 9 | 2.1 |
| Other | 6 | 1.4 |
| Locum Doctor | 4 | 0.9 |
| Specialist (SAS) | 4 | 0.9 |
| Professor | 1 | 0.2 |
| **Total** | **430** | **100** |

**Table 2.**
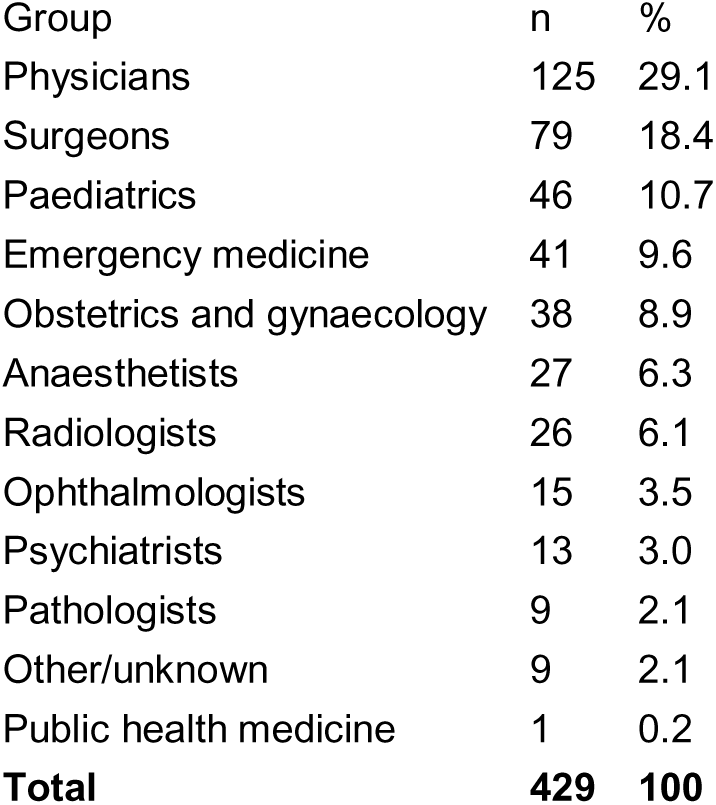
Specialty groupings of survey respondents (n = 429; 36 respondents did not report a specialty). See Table S1 for the full sub-specialty breakdown.

| Group | n | % |
| --- | --- | --- |
| Physicians | 125 | 29.1 |
| Surgeons | 79 | 18.4 |
| Paediatrics | 46 | 10.7 |
| Emergency medicine | 41 | 9.6 |
| Obstetrics and gynaecology | 38 | 8.9 |
| Anaesthetists | 27 | 6.3 |
| Radiologists | 26 | 6.1 |
| Ophthalmologists | 15 | 3.5 |
| Psychiatrists | 13 | 3.0 |
| Pathologists | 9 | 2.1 |
| Other/unknown | 9 | 2.1 |
| Public health medicine | 1 | 0.2 |
| **Total** | **429** | **100** |

### Patient complexity

Of those who completed the measure (n = 415), the average rating was 3.88 (median = 4; SD = 0.83; range = 1–5). The distribution of responses skewed toward higher complexity (Table S2).

### Mental workload (NASA-TLX)

Overall Raw-TLX scores were computed only for participants with complete data across all six subscales; participants with any missing subscale were excluded from the overall score but retained in item-level summaries. The pattern of results suggests that respondents’ elevated overall mental workload was driven primarily by cognitive and time-pressure burdens rather than by physical demand or frustration (Table 3).

**Table 3.** Means and standard deviations for the NASA-TLX Raw composite and six subscales.

| Measure | Mean | SD |
| --- | --- | --- |
| **Overall Raw-TLX (scaled 0–100)** | **66.2** | **11.5** |
| *Subscales (scaled 0–20)* |  |  |
| Mental demand | 16.0 | 2.9 |
| Temporal demand | 15.0 | 3.7 |
| Effort | 15.0 | 3.2 |
| Performance | 14.0 | 3.6 |
| Frustration | 11.7 | 5.0 |
| Physical demand | 7.0 | 5.0 |

### Interruptions and perceived impact

#### Self-reported frequency

A total of 391 respondents completed the interruptions measure. The average rating was 3.81 (SD = 0.99; median = 4; range = 1–5). Interruptions were a common feature of consultations: 61.1% of clinicians reported being interrupted often (30.7%) or very often (30.4%), and only one respondent (0.3%) reported never being interrupted (Table S3).

#### Perceived impact on mental workload and task resumption

A total of 391 participants responded to both items. The average rating for the perceived impact of interruptions on mental workload was 4.0 (SD = 1.11; median = 4; range = 1–5), indicating that interruptions were generally perceived as substantially increasing mental workload; 72.9% reported a high or significant impact (Table S4).

The average rating for the perceived difficulty of task resumption following an interruption was 3.15 (SD = 1.03; median = 3; range = 1–5), indicating that participants found resuming tasks moderately challenging on average. Resuming clinical tasks after interruptions was at least moderately challenging for three-quarters of clinicians (75.4% reporting levels 3–5), with 37.1% reporting that resumption was difficult or very difficult (Table S5).

#### Perceived impact on social factors

A total of 379 participants rated the perceived effect of interruptions on interactions with colleagues or other team members. The average rating was 2.74 (SD = 0.77; median = 3; range = 1–5), indicating a generally neutral-to-slightly negative perceived effect. Just over half of respondents (54.6%) rated the effect as neutral, 34.0% as negative or very negative, and 11.3% as positive or very positive (Table S6).

Framed as occurring during a typical clinic session, the perceived effect of interruptions on the interaction between the individual and a colleague in the moment was rated by 378 participants. The most common response (185 participants, 48.9%) was a negative impact on interactions with colleagues, with 159 (42.1%) reporting no significant impact and 34 (9.0%) a positive effect. Similarly, when framed as occurring across clinics and shifts, the perceived impact of interruptions on overall team cohesion was rated by 377 participants, with 169 (44.8%) reporting a negative impact, 163 (43.2%) no significant impact, and 45 (11.9%) a positive effect.

Taken together, these results indicate that interruptions were perceived to have a broadly negative impact on social and team dynamics. Approximately one third of participants reported a negative effect on their general interactions with colleagues, and close to half reported negative effects on both momentary collaboration during clinic sessions and overall team cohesion across shifts.

#### Self-reported strategies used

When asked about strategies used to manage interruptions, 146 of 377 participants (38.7%) reported adopting strategies, while 231 (61.3%) reported not doing so. Participants who reported using strategies were asked to describe them briefly; 145 provided usable text. For brevity, we report here on the sentiment and cluster analyses; details of the frequency analysis are in the Supplementary Materials (Tables S7 and S8).

#### Cluster analysis

The most interpretable solution emerging from *k*-means clustering used six-grams and a three-cluster model. The three clusters reflected distinct strategic orientations (exemplar quotes in Table 4):

1. **Cognitive offloading** — externalising tasks (e.g. asking others to wait, writing tasks down).
2. **Anticipating and preparing for interruptions** — proactive strategies (e.g. delegating to colleagues, avoiding interruption-prone spaces).
3. **Blunt responses / shutting down** — directly dismissing or blocking interruptions.

**Table 4.** Exemplar quotes for each cluster in the cluster analysis.

| Cluster | Sub-theme | Exemplar quote |
| --- | --- | --- |
| Cognitive offloading | Telling others to wait | “can it wait until I’ve finished the current clinical task” |
|  | Writing it down | “write information down as I go” |
| Anticipating and preparing for interruptions | Asking others to manage | “I ask in advance [...] ask a chaperone to deal with it” |
|  | Hiding to avoid interruptions | “try to hide in my room” |
| Blunt responses / shutting down | — | “often ends up being passive aggressive non verbal hostility” |

A chi-squared test showed that the distribution of NRC emotions differed significantly across clusters, χ²(14, N = 2133) = 174.4, p < .001. Blunt responses were characterised by higher anger, disgust and sadness, and lower anticipation and trust. Anticipating and preparing showed higher anticipation and trust, and lower anger, fear and disgust. Cognitive offloading was associated with higher surprise and fear, and lower anger. Figure S1 visualises the emotional profiles per cluster.

### Correlations of mental workload and predictors

Bivariate correlations between NASA-TLX outcomes and predictors are presented in Table 5. Overall workload (Raw-TLX) was most strongly associated with effort (*r* = .764), temporal demand (*r* = .745), and mental demand (*r* = .704). Among predictors, patient complexity, interruption frequency, and perceived cognitive impact showed the strongest and most consistent associations with workload across subscales. Perceived social effects of interruptions showed small negative associations with workload and frustration. Years of experience showed small effects, with more experienced doctors reporting slightly lower workload and better perceived performance. Strategy use was not consistently associated with workload.

**Table 5.** Bivariate Pearson correlations between NASA-TLX outcomes and study predictors.

| NASA-TLX outcome | Interruption freq. (n = 391) | Patient complexity (n = 401) | Cognitive impact (n = 391) | Resumption difficulty (n = 391) | Social effects (n = 379) | Strategies used (n = 377) | Years experience (n = 401) |
| --- | --- | --- | --- | --- | --- | --- | --- |
| Overall Raw-TLX | .343*** | .363*** | .393*** | .292*** | -.122* | .096 | -.064 |
| Mental demand | .251*** | .444*** | .331*** | .226*** | -.064 | .119* | -.080 |
| Temporal demand | .401*** | .237*** | .392*** | .330*** | -.096 | .060 | -.098* |
| Effort | .301*** | .375*** | .377*** | .291*** | -.112* | .137** | -.103* |
| Performance | -.127* | -.055 | -.162** | -.219*** | .182*** | -.004 | .186*** |
| Frustration | .238*** | .200*** | .355*** | .325*** | -.280**<br>* | .132** | -.101* |
| Physical demand | .166*** | .155** | .160** | .109* | .022 | -.057 | -.055 |

*Note.* \*\*\* $p < .001$ ; \*\* $p < .01$ ; \* $p < .05$ .

### Regression analyses

We evaluated the association between interruption frequency and perceived workload (NASA-TLX overall and each subscale) using sequential ordinary least-squares regression. For each workload outcome we fitted five nested models on the same complete-case sample: (1) interruption frequency only; (2) adding patient complexity and years of experience; (3) adding perceived cognitive impact and difficulty resuming tasks; (4) adding perceived team interaction effects; and (5) adding strategy use (binary). Full model coefficients are reported in Table S9.

#### Overall workload

Interruption frequency was associated with higher overall workload (unadjusted β = 4.23, SE = 0.57, p < .001) and remained significant in the fully adjusted model (β = 1.79, SE = 0.62, p = .004). However, much of this association was explained by patient complexity and the perceived cognitive impact of interruptions (Table S9; *R*² = 0.266).

#### Mental demand and effort

Similar patterns were observed for mental demand and effort. Interruption frequency appeared important in unadjusted analyses, but this association was attenuated by patient complexity and perceived cognitive impact; in fully adjusted models, neither mental demand (β = 0.18, SE = 0.16, p = .24) nor effort (β = 0.31, SE = 0.17, p = .071) retained significant independent associations with interruption frequency. For these dimensions of workload, the nature of the clinical work itself, compounded by how disruptive interruptions feel, appears to matter more than how often interruptions occur.

#### Physical demand

Physical demand showed a broadly similar pattern, though it was the weakest outcome overall (*R*² = 0.066). After accounting for patient complexity, the only significant predictor (β = 0.66, SE = 0.32, p = .041), interruption frequency was not a significant predictor of physical demand (β = 0.58, SE = 0.30, p = .057). This is intuitive given that more complex patients are likely to place greater demands on doctors regardless of interruptions.

#### Temporal demand

Temporal demand (the sense of working under time pressure) showed a different pattern. Interruption frequency remained a significant predictor (β = 0.94, SE = 0.20, p < .001) even after accounting for perceived cognitive impact (β = 0.68, p = .002) and patient complexity (β = 0.51, p = .018), both of which were also significant. This suggests that interruptions may impose a direct time cost distinct from their cognitive effects: they consume time that cannot be recovered.

#### Performance and frustration

For performance and frustration, the picture shifted again. Once other factors were accounted for, interruption frequency was no longer a significant predictor of either outcome. Instead, two other factors emerged as the dominant correlates. Difficulty resuming tasks after an interruption was independently associated with worse perceived performance (β = −0.59, SE = 0.24, p = .014), consistent with the idea that it is the effortful process of re-orienting to a clinical task, rather than the interruption itself, that undermines performance. The social experience of interruptions also mattered considerably: doctors who perceived interruptions as having a more positive effect on their interactions reported better perceived performance (β = 0.75, SE = 0.25, p = .003) and substantially lower frustration (β = −1.16, SE = 0.33, p < .001).

## Discussion

This study examined clinicians’ perceptions of interruption frequency and their cognitive and social correlates during secondary care outpatient consultations. Across a broad and professionally diverse sample, interruptions were perceived to be highly prevalent, with 61.1% of respondents reporting being interrupted often or very often during consultations. Interruption frequency, patient complexity, and the perceived cognitive impact of interruptions were each independently associated with higher mental workload across multiple NASA-TLX subscales. This study provides the first large-scale quantitative evidence of interruption prevalence specifically within secondary care outpatient practice in the UK.

The high prevalence of perceived interruptions extends existing evidence from primary care and ward settings to the outpatient secondary care context, where Shvartzman and Antonovsky [13] found 94% of primary care consultations were interrupted and qualitative evidence has consistently highlighted clinicians’ perceptions of interruptions as disruptive [14,15]. The elevated overall workload observed (mean Raw-TLX ≈ 66/100), particularly on the mental and temporal demand subscales, contextualises these findings against an already-pressured baseline, consistent with the prediction from CLT [5] that finite working memory capacity is routinely taxed in complex clinical settings and that any additional load, such as that imposed by an interruption, risks further degrading performance.

### Interruption frequency, patient complexity, and workload

Both interruption frequency and patient complexity were independently associated with higher workload across multiple subscales. Patient complexity was among the most consistent predictors, in line with frameworks identifying intrinsic cognitive load as a function of the number and interrelatedness of clinical elements [8,6,9]. The association between interruption frequency and workload was substantially attenuated after accounting for perceived cognitive impact, consistent with a process in which subjective cognitive burden, rather than occurrence alone, drives workload elevation. This aligns with evidence that an interruption’s disruptiveness depends on its timing, task complexity, and disruption to internal task representations [2,1], and suggests that reducing the cognitive cost of interruptions when they occur may be at least as important as reducing their frequency.

Task resumption difficulty was independently associated with poorer perceived performance even after controlling for interruption frequency and cognitive impact, identifying a discrete mechanism through which interruptions may undermine clinical work: the effortful re-orienting to an interrupted task. From a CLT perspective, this additional cognitive overhead, which involves reactivating interrupted task goals, retrieving working memory content, and re-establishing situational context, represents a form of extraneous load distinct from the interrupting event itself [7]. Interruption frequency nonetheless retained a direct association with temporal demand across all models, suggesting a distinct time-displacing effect not fully explained by cognitive burden, and consistent with evidence that interruptions extend consultation length and exacerbate time pressure [17]. Together, these findings are consistent with a dual-pathway model in which interruptions both *steal time*, raising temporal demand independently of other factors, and *impose cognitive costs* that, when perceived as burdensome, amplify effort and frustration.

### The limited protective effect of experience

We predicted that experience might buffer the negative effects of interruptions on mental workload, grounded in the expertise reversal hypothesis [11]. However, years of clinical experience showed only small and inconsistent associations with workload outcomes. More experienced clinicians reported marginally better perceived performance and slightly lower effort and frustration, but these effects largely diminished after adjustment for other predictors. This may reflect the fact that the patient complexity typical of secondary care settings exceeds the boundaries of automated processing even for experienced practitioners, or that the structural and environmental conditions that generate interruptions operate relatively independently of individual expertise. The implication is that interruption management cannot be delegated solely to individual skill development and must instead be addressed at system and team levels.

### Social dimensions of interruptions

Beyond their cognitive consequences, interruptions were perceived to have broadly negative social effects. Close to half of clinicians reported negative effects on immediate interactions with colleagues during clinic sessions, and a similar proportion reported negative effects on overall team cohesion across clinics and shifts. These findings are consistent with organisational research documenting that frequent interruptions can erode team communication norms and reduce interpersonal cohesion [18,19], and suggest that current interruption-management practices may be insufficient to carry the positive social benefits highlighted by Puranik et al. [20].

However, the perceived effect of interruptions on team interactions was one of the strongest predictors in the frustration and performance models. In the frustration model, a more negative perceived social impact of interruptions was associated with significantly higher frustration, even after accounting for cognitive impact and patient complexity. Conversely, in the performance model, perceiving interruptions as having a more positive effect on team interactions was associated with better perceived performance. This pattern suggests that the *social framing* of interruptions, whether they are experienced as disruptive or as constructive exchanges, may offset their downstream effects on clinician well-being and perceived performance. This is consistent with Puranik et al.’s [20] proposal that the interpersonal content of interruptions may partially offset their cognitive costs, and with evidence that brief, courteous exchanges during interruptions may sustain positive affect and social connection in a way that buffers frustration [18]. These findings underscore the value of approaches that preserve the communicative and relational functions of interruptions while minimising unnecessary cognitive disruption, for example through structured interprofessional communication frameworks such as ISBAR [21].

### Interruption-management strategies

Approximately 39% of clinicians reported using strategies to manage interruptions, yet strategy use was not associated with lower workload in any model. This mirrors the theoretical position of Puranik et al. [20] that the presence or absence of a strategy is unlikely to be informative, and extends it empirically: the cluster structure identified through NLP suggests that treating strategy use as binary conflates qualitatively distinct orientations. Anticipatory strategies reflect deliberate restructuring of workflow before interruptions occur; cognitive offloading manages their consequences by externalising task representations; blunt responses, characterised by higher anger and sadness, appear less like deliberate coping and more like expressions of accumulated frustration. Grouping these together in a binary variable likely masks heterogeneity that matters. It is plausible that proactive and offloading strategies do confer workload benefits, obscured in aggregate analyses by the inclusion of reactive, frustration-driven responses. This interpretation is tentative, being grounded in sentiment rather than direct measurement of intent.

We did not test whether cluster membership was differentially associated with workload; the corpus was too small (n = 145) to support such an analysis. The heterogeneity itself is nonetheless informative. It cautions against treating strategy use as a uniform construct in either research or intervention design, and points to strategy type as the more useful unit of analysis in future, adequately powered work. Future research should therefore measure strategy type rather than strategy presence, and evaluate whether proactive approaches are differentially effective across specialty and interruption type. The cluster structure identified here offers a theoretically grounded starting point for intervention design, targeting anticipatory planning and task resumption support at the team or system level rather than relying on individual improvisation.

### Implications for practice

Three practice-relevant findings emerge. First, because the association between interruption frequency and workload was substantially explained by perceived cognitive impact, interventions should address not only interruption frequency but also the cognitive cost of interruptions when they occur. Practical approaches include structured handover prompts, electronic task-bookmarking tools, and signalling norms that protect cognitively demanding work, drawing on the sterile cockpit principle from aviation [12,16]. Second, the finding that positive perceived social effects were associated with better performance and reduced frustration suggests that interventions should shape interpersonal exchanges to be purposeful and brief rather than eliminate them; structured communication frameworks such as ISBAR [21] may help preserve team-bonding functions while minimising cognitive disruption [20]. Third, because binary strategy use was unrelated to workload, effective strategies likely require co-design with clinicians and embedding at team and organisational levels rather than individual improvisation. Organisational approaches that protect uninterrupted time for complex consultations, such as longer scheduled slots, protected documentation time, or designated do-not-disturb periods, may offer particular benefit given the strength of patient complexity as a workload driver.

## Strengths and limitations

This study has several strengths. To our knowledge, it is the first national survey of interruptions in UK secondary care outpatient consultations, and the first to combine quantitative workload measurement with natural language processing of clinician-described strategies. The sample spans a wide range of specialties, grades (from SHO to consultant), and career stages (0–52 years post-qualification), providing a heterogeneous picture of contemporary outpatient practice. The NASA-TLX permitted decomposition of workload across cognitive, temporal, physical, and affective dimensions, and the sequential regression approach allowed systematic examination of attenuation across nested models. The NLP analysis of open-text strategy descriptions revealed heterogeneity that would have been invisible to binary coding alone.

Several limitations warrant acknowledgement. The cross-sectional design precludes causal inference; it is possible, for example, that clinicians experiencing high workload are more likely to perceive interruptions as frequent, rather than frequency driving workload. Recruitment was through a single medical defence organisation’s network and the eligible denominator could not be determined, as described under Sampling and Recruitment. The direction of any non-response bias is not self-evident: clinicians who find interruptions particularly disruptive may have been more motivated to respond, biasing prevalence estimates upward; equally, the busiest and most interruption-burdened clinicians may have been least able to take time to complete the survey, biasing estimates downward. The predominance of consultants (75.8% of those reporting a role) and the mean of 18.9 years since qualification indicate that the sample skews towards senior clinicians, and findings may generalise less well to trainees. The use of single-item Likert measures for several constructs was a deliberate trade-off to reduce respondent burden and maximise completion in a high-workload population, but it limits measurement precision and precludes conventional internal-consistency reliability testing; future work using validated multi-item instruments would strengthen confidence in these estimates. Binary coding of strategy use oversimplifies what the NLP reveals as richly varied practice. Finally, the absence of objective interruption data, patient outcomes, or environmental moderators (office layout, electronic health record system, clinic staffing) means that relationships between perceived workload and patient safety remain unestablished.

Future research should combine self-report with observational, time-motion, and EHR-derived methods; develop and rigorously evaluate targeted interventions using randomised or stepped-wedge designs; and use qualitative and longitudinal approaches to examine contextual moderators and adaptation over time. Formal mediation models, and tests of interaction between strategy type and interruption context, are logical next steps in adequately powered samples.

## Conclusion

Interruptions are a routine feature of secondary care outpatient practice and are associated with higher perceived workload, with much of this association explained by clinical complexity and the subjective cognitive burden of interruptions. Interruption frequency nonetheless retained a direct association with temporal pressure after adjustment, indicating that time pressure is a distinct consequence independent of cognitive burden. Workload reduction efforts should therefore address both interruption frequency and cognitive cost, while supporting task resumption and constructive team interaction in context-sensitive ways.

## Supporting information

Supplementary Materials referenced in text

## Data Availability

Data are held within secured UCL networks, accessible only to the named research team, in accordance with the terms of the ethical approval, and will be retained for no more than 10 years.

## Declarations

### Contributors

SB conceived and designed the study, obtained ethical approval and funding, led data collection, conducted the statistical and natural language processing analyses, and drafted the manuscript. CS contributed to data collection and data analysis, and to drafting the manuscript. AR contributed to the conceptualisation and design of the study and critically revised the manuscript for important intellectual content. AG contributed to the conceptualisation and design of the study and critically revised the manuscript for important intellectual content. All authors approved the final version of the manuscript and agree to be accountable for all aspects of the work. SB is the guarantor and accepts full responsibility for the work and the conduct of the study, had access to the data, and controlled the decision to publish.

### Patient consent for publication

Not applicable.

### Ethics approval

This study involved human participants and was approved by the UCL Research Ethics Committee on 20 May 2025 (ID: 1132). All participants provided informed consent before taking part.

### Data availability statement

Data are not publicly available. Under the terms of the UCL research ethics approval (ID: 1132), survey data are held within the UCL Data Safe Haven and are accessible only to the named research team. The R code used for the analyses reported here is available from the corresponding author on reasonable request.

### Provenance and peer review

Not commissioned; externally peer reviewed.

### Supplementary material

Supplementary Materials (Supplements 1–3, Tables S1–S9, Figure S1) and the analysed survey items are provided as separate files.

