## Supplementary Materials referenced in text for "Doctors’ experiences of interruptions in UK secondary care outpatient clinics: a cross-sectional survey of prevalence, consequences and management strategies"

### SUPPLEMENT 1: DETAILS OF MEASURES

**Patient Complexity.** Participants rated their typical patient complexity using a single item on a five-point scale (1 = *Lowest complexity* to 5 = *Highest complexity*). Patient complexity was used as a proxy for intrinsic task demand, consistent with frameworks in which the number and interrelatedness of clinical elements determine cognitive load (Liu & Li, 2012; Nicolaus et al., 2022).

**Mental Workload.** Mental workload was assessed using the NASA Task Load Index (NASA-TLX; Hart et al., 1988), adapted for use in healthcare settings (Hoonakker et al., 2011; Huggins & Claudio, 2018; Tubbs-Cooley et al., 2018). Participants rated six subscales on 0–20 scales: Mental Demand, Physical Demand, Temporal Demand, Performance, Effort, and Frustration. An overall Raw-TLX score was computed as the unweighted arithmetic mean of all six subscales, scaled to 0–100 (Raw-TLX × 5). Participants with any missing subscale data were excluded from the overall Raw-TLX score but retained in subscale-level analyses. Following common practice in healthcare NASA-TLX adaptations, the Performance subscale was reverse-scored prior to analysis so that higher values indicate better self-rated performance; this scoring is preserved throughout the Results and Tables.

**Interruption Frequency.** Self-reported interruption frequency during consultations was assessed with a single item rated on a five-point Likert scale (1 = *Never* to 5 = *Very often*; see Yu & Lee, 2022).

**Perceived Cognitive Impact of Interruptions.** The perceived impact of interruptions on mental workload was assessed using a single item rated on a five-point Likert scale (1 = *No impact* to 5 = *Significant impact*).

**Task Resumption Difficulty.** The perceived difficulty of resuming a task following an interruption was assessed using a single item rated on a five-point Likert scale (1 = *Very easy* to 5 = *Very difficult*).

**Social Effects of Interruptions.** No validated self-report scale currently exists for assessing the perceived social consequences of interruptions in clinical settings. Items were therefore developed for this study, drawing on theoretical frameworks identifying two separable social dimensions of interruptions: their immediate interpersonal impact during an interaction, and their broader effect on team cohesion over time (Puranik et al., 2020, 2021; Toebben et al., 2024). The first used a five-point scale (1 = *Very negative* to 5 = *Very positive*) to capture the general perceived effect of interruptions on interactions with colleagues or team members. Two further items used three-point scales (*Negatively*, *No impact*, *Positively*) to assess the perceived impact of interruptions on immediate colleague interactions during a typical clinic session, and on overall team cohesion across clinics and shifts.

**Interruption-Management Strategy Use.** Participants were asked to report whether they used any formal or informal strategies to manage interruptions (yes/no). Those who responded affirmatively were invited to describe their strategy in an open-text field. Strategy use was coded as a binary variable for quantitative analyses.

### SUPPLEMENT 2: ADDITIONAL TABLES

#### Table S1. Detailed breakdown of specialty (n = 429; 36 respondents did not report a specialty). Percentages are of the 429 respondents reporting a specialty and may not sum to 100 owing to rounding.

| **Group** | **Specialty** | **N** | **Proportion %** |
| --- | --- | --- | --- |
| **Physicians** |  | ***125*** | ***29.1*** |
|  | Acute internal medicine | 13 | 3.0 |
|  | Cardiology | 9 | 2.1 |
|  | Clinical genetics | 2 | 0.5 |
|  | Clinical pharmacology and therapeutics | 1 | 0.2 |
|  | Dermatology | 11 | 2.6 |
|  | Endocrinology and diabetes mellitus | 10 | 2.3 |
|  | Gastro-enterology | 10 | 2.3 |
|  | General (internal) medicine | 3 | 0.7 |
|  | Genito-urinary medicine | 6 | 1.4 |
|  | Geriatric medicine | 15 | 3.5 |
|  | Medical oncology | 3 | 0.7 |
|  | Neurology | 6 | 1.4 |
|  | Palliative medicine | 7 | 1.6 |
|  | Renal medicine | 4 | 0.9 |
|  | Respiratory medicine | 15 | 3.5 |
|  | Rheumatology | 9 | 2.1 |
|  | Tropical medicine | 1 | 0.2 |
| **Surgeons** |  | ***79*** | ***18.4*** |
|  | Cardio-thoracic surgery | 2 | 0.5 |
|  | General surgery | 23 | 5.4 |
|  | Neurosurgery | 3 | 0.7 |
|  | Oral and maxillo-facial surgery | 3 | 0.7 |
|  | Otolaryngology | 6 | 1.4 |
|  | Plastic surgery | 5 | 1.2 |
|  | Trauma and orthopaedic surgery | 25 | 5.8 |
|  | Urology | 11 | 2.6 |
|  | Vascular surgery | 1 | 0.2 |
| **Radiologists** |  | ***26*** | ***6.1*** |
|  | Clinical oncology | 8 | 1.9 |
|  | Clinical radiology | 18 | 4.2 |
| **Pathologists** |  | ***9*** | ***2.1*** |
|  | Chemical pathology | 2 | 0.5 |
|  | Haematology | 6 | 1.4 |
|  | Histopathology | 1 | 0.2 |
| **Psychiatrists** |  | ***13*** | ***3.0*** |
|  | Child and adolescent psychiatry | 2 | 0.5 |
|  | Forensic psychiatry | 1 | 0.2 |
|  | General psychiatry | 7 | 1.6 |
|  | Old age psychiatry | 2 | 0.5 |
|  | Psychiatry of learning disability | 1 | 0.2 |
| **Paediatrics** |  | ***46*** | ***10.7*** |
|  | Paediatric cardiology | 1 | 0.2 |
|  | Paediatrics | 45 | 10.5 |
| **Obstetrics and gynaecology** |  | ***38*** | ***8.9*** |
| **Ophthalmologists** |  | 15 | 3.5 |
| **Anaesthetists** |  | 27 | 6.3 |
| **Emergency Medicine** |  | 41 | 9.6 |
| **Public health medicine** |  | 1 | 0.2 |
| **Other/Unknown** | **Group total** | 9 | 2.1 |
| **Other/Unknown** | **Other** | 8 | 1.9 |
| **Other/Unknown** | **Rehabilitation medicine** | 1 | 0.2 |

#### Table S2. Distribution of self-reported patient complexity profile.

|  | Count (n) | Percent % |
| --- | --- | --- |
| Highest complexity | 105 | 25.3% |
| Second highest | 169 | 41.0% |
| Mid-point | 130 | 31.3% |
| Low | 8 | 1.9% |
| Lowest | 3 | 0.7% |

#### Table S3. Distribution of self-reported interruption frequency.

|  | Count (n) | Percent |
| --- | --- | --- |
| Very often | 119 | 30.4% |
| Often | 120 | 30.7% |
| Sometimes | 111 | 28.4% |
| Rarely | 40 | 10.2% |
| Never | 1 | 0.26% |

#### Table S4. Distribution of self-reported impact on mental workload.

| Mental Workload | Count (n) | Percent |
| --- | --- | --- |
| Significant impact | 167 | 42.7% |
| High impact | 118 | 30.2% |
| Moderate impact | 57 | 14.6% |
| Mild impact | 37 | 9.5% |
| No impact | 12 | 3.1% |

#### Table S5. Distribution of self-reported impact on task resumption.

| Task Resumption | Count (n) | Percent |
| --- | --- | --- |
| Very difficult | 34 | 8.7% |
| Difficult | 111 | 28.4% |
| Moderately challenging | 150 | 38.4% |
| Easy | 70 | 17.9% |
| Very easy | 26 | 6.7% |

#### Table S6. Distribution of self-reported impact on interaction with colleagues.

| Interactions with Colleagues | Count (n) | Percent |
| --- | --- | --- |
| Very negative | 18 | 4.8% |
| Negative | 111 | 29.3% |
| Neutral / Midpoint | 207 | 54.6% |
| Positive | 36 | 9.5% |
| Very positive | 7 | 1.9% |

### SUPPLEMENT 3: ANALYSIS OF INTERRUPTION STRATEGIES USED

#### Word Frequency Analysis

To establish a descriptive overview of the data, we quantified the frequency of individual words and higher-order n-grams (bigrams, trigrams, six-grams; i.e., sequences of words). N-gram analysis was used to identify frequently co-occurring terms and recurring phrases that provide more context than single words. See Table S7.

**Table S7.**
*Exemplar frequent uni-grams, tri-grams, six-grams.*

| Uni-Grams | n | Tri-Grams | n | Six-Grams | n |
| --- | --- | --- | --- | --- | --- |
| Wait | 27 | to wait until | 9 | to wait until I have finished | 3 |
| Phone | 25 | wait until I | 8 | I ask not to be interrupted | 2 |
| Clinic | 24 | I have finished | 6 | I might ask someone to hold | 2 |
| Patient | 20 | until I have | 6 | might ask someone to hold my | 2 |
| Task | 19 | ask people to | 5 | I briefly assess the urgency severity | 1 |
| Urgent | 19 | ask them to | 5 | 1. Delegate tasks. 2. Delay action. | 1 |

#### Sentiment Analysis

Sentiment analysis using a standard semantic word base (the Bing Lexicon; Hu & Liu, 2004), and accounting for simple negations (e.g., not happy, never helpful), categorised all 145 strategies reported as negatively valanced, and 53 as positively valanced, indicating that 1/3 of strategies reflected both positive and negative approaches to interruptions and their management.

The *NRC Emotion Lexicon* (Mohammad & Turney, 2013) was then applied to classify responses according to eight basic emotions (anger, anticipation, disgust, fear, joy, sadness, surprise, and trust; Plutchik, 1980). Anticipation appeared in nearly all responses, alongside frequent mentions of trust, fear, and surprise, suggesting preparation for interruptions and reliance on others, while still experiencing them as unexpected. See Table S8.

**Table S8.**
*Number of Responses Containing Each NRC Emotion.*

| Sentiment | Sentiment Description | N |
| --- | --- | --- |
| Anticipation | Expecting, preparing for, or worrying about future interruptions. | 86 |
| Trust | Reliance on colleagues, processes, or strategies to handle interruptions. | 55 |
| Fear | Anxiety or concern about the impact of interruptions. | 51 |
| Surprise | Reactions to interruptions that are unexpected. | 47 |
| Sadness | Disappointment or discouragement linked to interruptions. | 42 |
| Joy | Satisfaction when strategies to manage interruptions are effective. | 21 |
| Anger | Frustration or irritation caused by interruptions. | 20 |
| Disgust | Strong aversion or rejection of interruptions. | 11 |

*Note. 1.* Sentiment descriptions have been adapted to suit the context of the present research while aligning with the descriptions provided by Mohammad & Turney (2013) and Plutchik (1980). 2. Each response was often characterised by multiple emotions, indicated by a higher number of total emotions than total participants.

#### Table S9. Analysis of Strategies Used

|  | Model 1 | Model 2 | Model 3 | Model 4 | Model 5 |
| --- | --- | --- | --- | --- | --- |
| NASA-TLX - Overall workload |  |  |  |  |  |
| Interruption Frequency | 4.23 (0.57)*** | 3.27 (0.56)*** | 1.82 (0.62)** | 1.81 (0.62)** | 1.79 (0.62)** |
| Patient Complexity | — | 4.17 (0.68)*** | 3.69 (0.67)*** | 3.69 (0.67)*** | 3.70 (0.67)*** |
| Years of Experience | — | −0.09 (0.05) | −0.06 (0.05) | −0.06 (0.05) | −0.06 (0.05) |
| Perceived Impact on Cognitive Load | — | — | 2.64 (0.68)*** | 2.69 (0.69)*** | 2.67 (0.69)*** |
| Difficulty Resuming Tasks | — | — | 0.21 (0.70) | 0.29 (0.71) | 0.29 (0.71) |
| Impact on Team Interactions | — | — | — | 0.49 (0.73) | 0.49 (0.73) |
| Strategy Use | — | — | — | — | 0.28 (1.11) |
| Model R² | **0.13** | **0.215** | **0.265** | **0.266** | **0.266** |
| Mental demands |  |  |  |  |  |
| Interruption Frequency | 0.82 (0.15)*** | 0.49 (0.14)*** | 0.22 (0.16) | 0.21 (0.16) | 0.18 (0.16) |
| Patient Complexity | — | 1.44 (0.17)*** | 1.35 (0.17)*** | 1.35 (0.17)*** | 1.36 (0.17)*** |
| Years of Experience | — | −0.02 (0.01) | −0.01 (0.01) | −0.01 (0.01) | −0.02 (0.01) |
| Perceived Impact on Cognitive Load | — | — | 0.60 (0.17)*** | 0.61 (0.17)*** | 0.59 (0.17)*** |
| Difficulty Resuming Tasks | — | — | −0.10 (0.17) | −0.07 (0.18) | −0.07 (0.18) |
| Impact on Team Interactions | — | — | — | 0.18 (0.18) | 0.19 (0.18) |
| Strategy Use | — | — | — | — | 0.41 (0.28) |
| Model R² | **0.077** | **0.232** | **0.263** | **0.265** | **0.269** |
| Physical demands |  |  |  |  |  |
| Interruption Frequency | 0.85 (0.26)** | 0.68 (0.26)* | 0.53 (0.30) | 0.51 (0.30) | 0.58 (0.30) |
| Patient Complexity | — | 0.73 (0.32)* | 0.69 (0.32)* | 0.69 (0.32)* | 0.66 (0.32)* |
| Years of Experience | — | −0.04 (0.03) | −0.04 (0.03) | −0.04 (0.03) | −0.03 (0.03) |
| Perceived Impact on Cognitive Load | — | — | 0.38 (0.33) | 0.44 (0.33) | 0.49 (0.33) |
| Difficulty Resuming Tasks | — | — | −0.13 (0.34) | −0.03 (0.34) | −0.01 (0.34) |
| Impact on Team Interactions | — | — | — | 0.59 (0.35) | 0.57 (0.35) |
| Strategy Use | — | — | — | — | −0.91 (0.54) |
| Model R² | **0.028** | **0.048** | **0.052** | **0.059** | **0.066** |
| Temporal demands |  |  |  |  |  |
| Interruption Frequency | 1.52 (0.17)*** | 1.37 (0.18)*** | 0.93 (0.20)*** | 0.93 (0.20)*** | 0.94 (0.20)*** |
| Patient Complexity | — | 0.67 (0.22)** | 0.51 (0.21)* | 0.51 (0.21)* | 0.51 (0.21)* |
| Years of Experience | — | −0.03 (0.02) | −0.02 (0.02) | −0.02 (0.02) | −0.02 (0.02) |
| Perceived Impact on Cognitive Load | — | — | 0.65 (0.22)** | 0.67 (0.22)** | 0.68 (0.22)** |
| Difficulty Resuming Tasks | — | — | 0.24 (0.22) | 0.28 (0.23) | 0.28 (0.23) |
| Impact on Team Interactions | — | — | — | 0.18 (0.23) | 0.18 (0.23) |
| Strategy Use | — | — | — | — | −0.24 (0.36) |
| Model R² | **0.17** | **0.2** | **0.24** | **0.24** | **0.24** |
| Effort |  |  |  |  |  |
| Interruption Frequency | 1.05 (0.16)*** | 0.76 (0.15)*** | 0.34 (0.17)* | 0.33 (0.17)* | 0.31 (0.17) |
| Patient Complexity | — | 1.27 (0.19)*** | 1.13 (0.18)*** | 1.12 (0.18)*** | 1.13 (0.18)*** |
| Years of Experience | — | −0.03 (0.01)* | −0.03 (0.01) | −0.03 (0.01) | −0.03 (0.01) |
| Perceived Impact on Cognitive Load | — | — | 0.68 (0.19)*** | 0.69 (0.19)*** | 0.67 (0.19)*** |
| Difficulty Resuming Tasks | — | — | 0.17 (0.19) | 0.18 (0.19) | 0.18 (0.19) |
| Impact on Team Interactions | — | — | — | 0.07 (0.20) | 0.08 (0.20) |
| Strategy Use | — | — | — | — | 0.36 (0.31) |
| Model R² | **0.106** | **0.21** | **—** | **—** | **0.271** |
| Performance |  |  |  |  |  |
| Interruption Frequency | −0.50 (0.19)** | −0.49 (0.19)* | −0.19 (0.21) | −0.21 (0.21) | −0.23 (0.21) |
| Patient Complexity | — | −0.05 (0.23) | 0.07 (0.23) | 0.07 (0.23) | 0.07 (0.23) |
| Years of Experience | — | 0.07 (0.02)*** | 0.06 (0.02)*** | 0.06 (0.02)*** | 0.06 (0.02)** |
| Perceived Impact on Cognitive Load | — | — | −0.03 (0.24) | 0.05 (0.23) | 0.04 (0.23) |
| Difficulty Resuming Tasks | — | — | −0.71 (0.24)** | −0.59 (0.24)* | −0.59 (0.24)* |
| Impact on Team Interactions | — | — | — | 0.74 (0.25)** | 0.75 (0.25)** |
| Strategy Use | — | — | — | — | 0.24 (0.38) |
| Model R² | **0.019** | **0.054** | **0.089** | **0.11** | **0.11** |
| Frustration |  |  |  |  |  |
| Interruption Frequency | 1.34 (0.25)*** | 1.12 (0.26)*** | 0.36 (0.28) | 0.39 (0.28) | 0.36 (0.28) |
| Patient Complexity | — | 0.96 (0.31)** | 0.68 (0.31)* | 0.68 (0.30)* | 0.70 (0.30)* |
| Years of Experience | — | −0.05 (0.02)* | −0.04 (0.02) | −0.03 (0.02) | −0.04 (0.02) |
| Perceived Impact on Cognitive Load | — | — | 0.88 (0.31)** | 0.76 (0.31)* | 0.74 (0.31)* |
| Difficulty Resuming Tasks | — | — | 0.77 (0.32)* | 0.57 (0.32) | 0.57 (0.32) |
| Impact on Team Interactions | — | — | — | −1.18 (0.33)*** | −1.16 (0.33)*** |
| Strategy Use | — | — | — | — | 0.48 (0.50) |
| Model R² | **0.07** | **0.1** | **0.18** | **0.2** | **0.21** |
| *Legend:* SE in parentheses. *** p < .001, ** p < .01, * p < .05. — variable not included in that model. | | | | | |

#### Figure S1. Distribution of Emotions per Cluster.

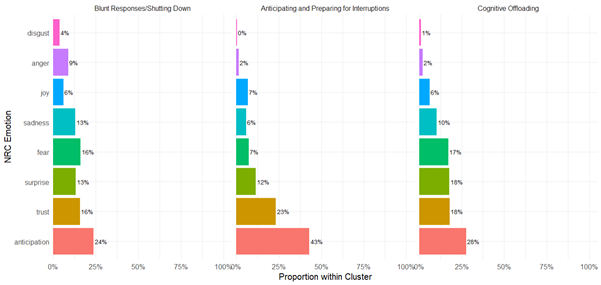
